# Generative AI for Thematic Analysis in a Maternal Health Study: Coding Semi-structured Interviews using Large Language Models (LLMs)

**DOI:** 10.1101/2024.09.16.24313707

**Authors:** Shan Qiao, Xingyu Fang, Junbo Wang, Ran Zhang, Xiaoming Li, Yuhao Kang

**Affiliations:** Department of Health Promotion, Education, and Behavior, Arnold School of Public Health, University of South Carolina; GISense Lab, Department of Geography and the Environment, The University of Texas at Austin; Department of Geography, University of South Carolina; School of Civil, Environmental and Geomatic Engineering, University College London, London, United Kingdom; Department of Geography and Sustainability, University of Tennessee, Knoxville

## Abstract

**Study Objectives:** The coding of semi-structured interview transcripts is a critical step for thematic analysis of qualitative data. However, the coding process is often labor-intensive and time-consuming. The emergence of generative artificial intelligence (GenAI) presents new opportunities to enhance the efficiency of qualitative coding. This study proposed a computational pipeline using GenAI to automatically extract themes from interview transcripts.

**Methods:** Using transcripts from interviews conducted with maternity care providers in South Carolina, we leveraged ChatGPT for inductive coding to generate codes from interview transcripts without a predetermined coding scheme. Structured prompts were designed to instruct ChatGPT to generate and summarize codes. The performance of GenAI was evaluated by comparing the AI-generated codes with those generated manually.

**Results:** GenAI demonstrated promise in detecting and summarizing codes from interview transcripts. ChatGPT exhibited an overall accuracy exceeding 80% in inductive coding. More impressively, GenAI reduced the time required for coding by 81%.

**Discussion:** GenAI models are capable of efficiently processing language datasets and performing multi-level semantic identification. However, challenges such as inaccuracy, systematic biases, and privacy concerns must be acknowledged and addressed. Future research should focus on refining these models to enhance reliability and address inherent limitations associated with their application in qualitative research.

## 1. Introduction

Qualitative research commonly involves collecting and analyzing non-numerical data (e.g., text, video, audio) to understand concepts, perceptions, beliefs, attitudes, opinions, and lived experiences within specific contexts [1], [2]. Qualitative studies have been widely used in social sciences and public health research to generate in-depth insights, discover latent patterns, and inform the development of new research directions [3], [4], [5]. In mixed-methods research, qualitative methods serve both exploratory and explanatory functions, uncovering salient constructs for quantitative measurement and providing detailed interpretations of quantitative results [6], [7], [8]. The choice of a qualitative research approach by researchers depends on the theoretical tradition and research paradigm that they adhere to.

As a form of inquiry, qualitative research is diverse and consists of many research paradigms that shape the assumptions a researcher adheres to when answering their research question(s) of interest [9]. For example, postpositivism as a research paradigm adheres to the worldview that even though there is a true reality, the ability to know everything about it is impossible, as there is always the possibility of new knowledge arising that disproves existing knowledge [9], [10]. Epistemologically, postpositivists believe that knowledge is valid when it is generated by other researchers and scientific experts. Conversely, constructivism as a research paradigm assumes that multiple realities exist [9], [10]. Rooted in hermeneutics, constructivism assumes that these realities are constructed within each individual’s experience and can be uncovered through rich discussion between researchers and study participants [11]. Unlike postpostivsim, constructism epistemologically assumes that knowledge generation is a co-constructive process between researchers and study participants [11]. In a separate vein, the critical theory research paradigm stems from social movements and the Frankfurt School [12], [13]. Ontologically, the critical theory paradigm assumes that an individual’s reality is shaped by socio-contextual factors (i.e., social, political gender, cultural, and race/ethnicity) [10], [12]. Epistemologically, the critical theory paradigm assumes that knowledge is cooperatively produced between researchers and the populations featured within their studies [12], [13]. Regardless of which paradigm is selected by a researcher, adhering to their underlying assumptions is important, as they provide the underlying rationale to assess the reasonableness of a study’s methods, analyses, and conclusions [9], [10], [11], [12].

Thematic analysis can be used to analyze patterns across the respondents within a study sample, as outlined in a six-step process by Braun and Clarke [14]. It includes familiarizing, generating initial codes, searching for themes, reviewing themes, defining and naming themes, and producing the report. Within Braun and Clarke’s approach to thematic analysis, interviews are one type of data collection strategy in which trained interviewers ask participants questions based on interview guides that cover key topics [14], [15]. Depending on the research team’s onto-epistemology and qualitative research tradition, interviews can be structured, semi-structured, or unstructured [16]. The process of analyzing interview data within this analytical approach typically requires researchers to transcribe audio recordings verbatim, review the transcripts, code the data, and group these codes into latent level themes. Within thematic analysis, coding is not only a critical step in the data analysis process but it also essential for getting familiarized with the data [14].

In thematic analysis more generally, coding is a process of assigning labels to data (e.g., words, phrases, sentences, or paragraphs) that in relation to a research question of interest [17], [18]. There are many different approaches to coding qualitative data, two approaches include an inductive approach and a deductive approach [19]. Deductive approaches to coding are often theory-driven, consisting of a predefined coding scheme (e.g., based on theory or literature review) that sorts data into preset categories [20]. It relies on existing theories, concepts, or frameworks to guide the coding process [19], [20]. Inductive approaches to coding do not use a predefined coding scheme but instead involve organically generating codes in relation to a research question [19], [21].

Coding interviews is a time-intensive process that involves an in-depth reading and understanding of each transcript. Depending on the decided unit of analysis (e.g. single lines of text, paragraphs, a complete document like a transcript) the amount of time needed to code an interview transcript can vary greatly [22]. In recent years, with the development of computer software, many qualitative data analysis software have begun to provide coding functions, helping researchers efficiently manage and organize their qualitative datasets [23]. However, the cost of expensive software and the opaque data processing workflow may be challenges for individual researchers or small research teams with limited budgets.

The emergence of Generative Artificial Intelligence (GenAI), with the development of computer science tools and methods, offers promising opportunities for qualitative research [24]. The field of Natural Language Processing (NLP) has made significant advancements in enabling computers to understand human languages and manipulate texts [25]. A notable example of this advancement refers to ChatGPT, a cutting-edge NLP model developed by OpenAI [26]. ChatGPT is based on the Generative Pre-trained Transformer (GPT) model, which utilizes the Transformer architecture to generate coherent and contextually appropriate responses in human-like conversational settings [27]. By training on vast amounts of text data, ChatGPT can generate coherent, contextually relevant responses to a wide range of human language inputs. More importantly, the superior ability of ChatGPT to understand language allows it to engage in dynamic and natural conversations. This deep understanding enables ChatGPT to respond in ways that are contextually appropriate, nuanced, and reflective of human-like reasoning, creating opportunities across a wide range of domains.

Researchers are increasingly exploring the integration of ChatGPT into various fields, recognizing its potential to revolutionize the handling of qualitative data. For instance, researchers in psychology, education, and public health have begun investigating how ChatGPT can be leveraged to facilitate their tasks such as deductive coding, inductive coding, and thematic analysis [28], [29], [30], [31], [32], [33]. These pioneering efforts highlight the promise of LLMs in enhancing the efficiency and scalability of qualitative research. Despite their success, prior research has also noted that LLMs, including ChatGPT, often lack domain-specific knowledge [34]. There has been limited exploration of the applications of LLMs to support maternal health studies, highlighting the need for interdisciplinary collaboration to incorporate advanced technology with real-world practices.

To this end, our study examines the potential of ChatGPT, in particular, ChatGPT 4 model, to assist with the inductive coding process for thematic analysis in qualitative studies, specifically following Braun and Clarke’s widely adopted framework [14]. Using semi-structured interview data collected from maternity care providers in South Carolina, this study (1) proposes a novel computational workflow to use ChatGPT in inductive coding of thematic analysis, as well as (2) assesses the coding performance of ChatGPT in terms of its credibility and dependability, by comparing its coding results with those coded manually. Notably, OpenAI offers the access to ChatGPT through an Application Programming Interface (API), which supports the automated processing of multiple input documents. Thus, we will leverage the ChatGPT API into our workflow to support thematic analysis and generate codes.

This study is situated within a postpositivist research paradigm. Therefore, this study was conducted with the ontological assumption that while a single reality exists, it may not be possible to understand it in its entirety since there are factors that we cannot account for [9], [10]. Likewise, this study epistemologically assumes that elements of quantitative forms of inquiry (e.g. statistics) can be used if needed to answer the study’s research question [9], [10]. Moreover, this study epistemologically believes that other researchers are viable assessors of the validity of our work [9], [10]. We chose to situate this study within a postpositivist research paradigm because it allows us to flexibly conceptualize how to apply quantitative technology developed using positivist rationale (i.e., GenAI) within a qualitative research context. This study could contribute to this emerging area by illustrating how LLMs can be leveraged to enhance qualitative analysis in maternal health research, with potential implications for broader applications across public health domains.

## 2. Methods

### 2.1. Interview Raw Data

The qualitative data used in this study were derived from a previous qualitative study that investigated racial disparities in maternal healthcare services and outcomes during the COVID-19 pandemic in South Carolina. In that study, we conducted semi-structured interviews with 39 women who gave birth between March 2020 and July 2021 and 9 maternity care providers (i.e., physicians, nurses, and case managers) from clinics serving communities with a high proportion of Black and/or Hispanic populations. In the current study, only the data from the maternity care providers was used. More details about the dataset and study protocol can be referenced in Zhang et al. [35]. The audio recordings of the interviews were transcribed using Otter.ai [36], and the transcripts were manually reviewed and refined by two research assistants to ensure accuracy. The interview guide questions are listed in Table 1.

**Table 1.**
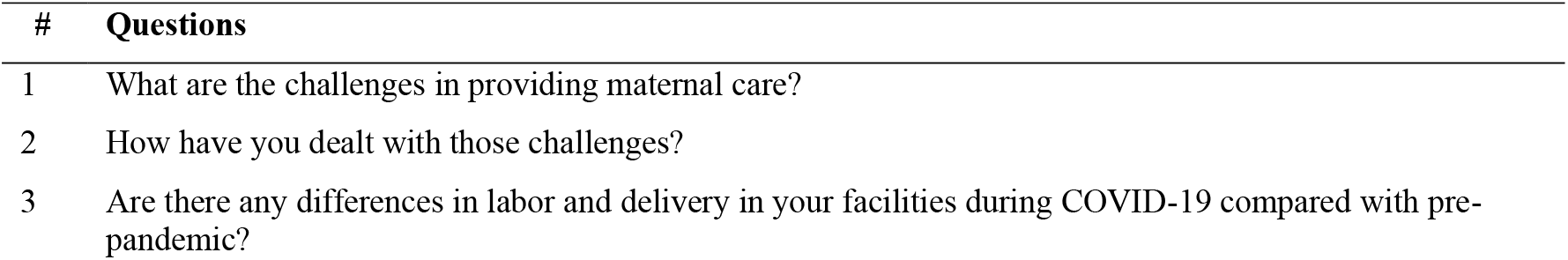
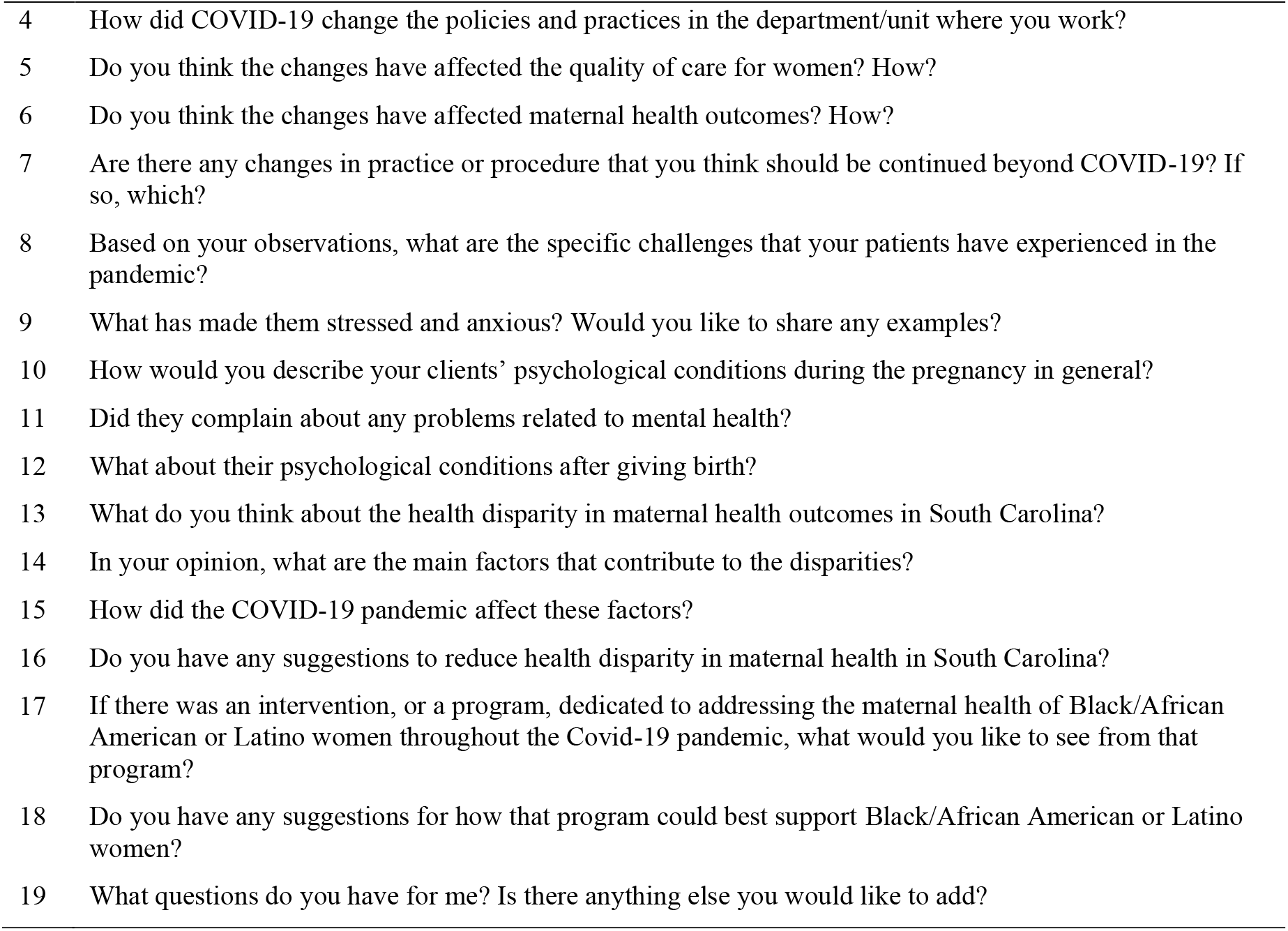
Questions of the in-depth interview guide from the maternity care providers.

### 2.2 Research Ethics of Reusing Data

All personal and identifiable information was removed from the transcripts to protect participants’ privacy and confidentiality before conducting data analysis, following our standardized qualitative study protocol [35]. In addition, we applied for and obtained Institutional Review Board (IRB) approval for an amendment allowing the reuse of transcripts collected for a traditional qualitative study for use in the present context. Considering potential ethical concerns and privacy protection, we chose to exclude the transcripts or data from ethnic minority women in this exploratory study. The reuse of the transcript data was approved as a study amendment by the IRB committee at the University of South Carolina (#Pro00115169).

### 2.3 Interview Transcript and Structure

Given the structure of an interview, the raw interview transcript typically follows a question-and- answer structure that allows researchers to consider a variety of responses from different participants to the same question, thereby identifying diverse opinions and lived experiences. To facilitate GenAI’s coding, we first reformatted the text into structured, analyzable data by matching each question with its corresponding answer. Each question-answer pair (hereafter referred to as a *dialogue*) was assigned a unique identifier (ID#), making the data computer-readable and facilitating efficient data management and query. Figure 1 illustrates the reformatting process from raw text into structured data, where interviewers’ questions were aligned with participants’ responses. This structured data was saved in .CSV files, with each participant’s responses organized accordingly.

**Figure 1.**
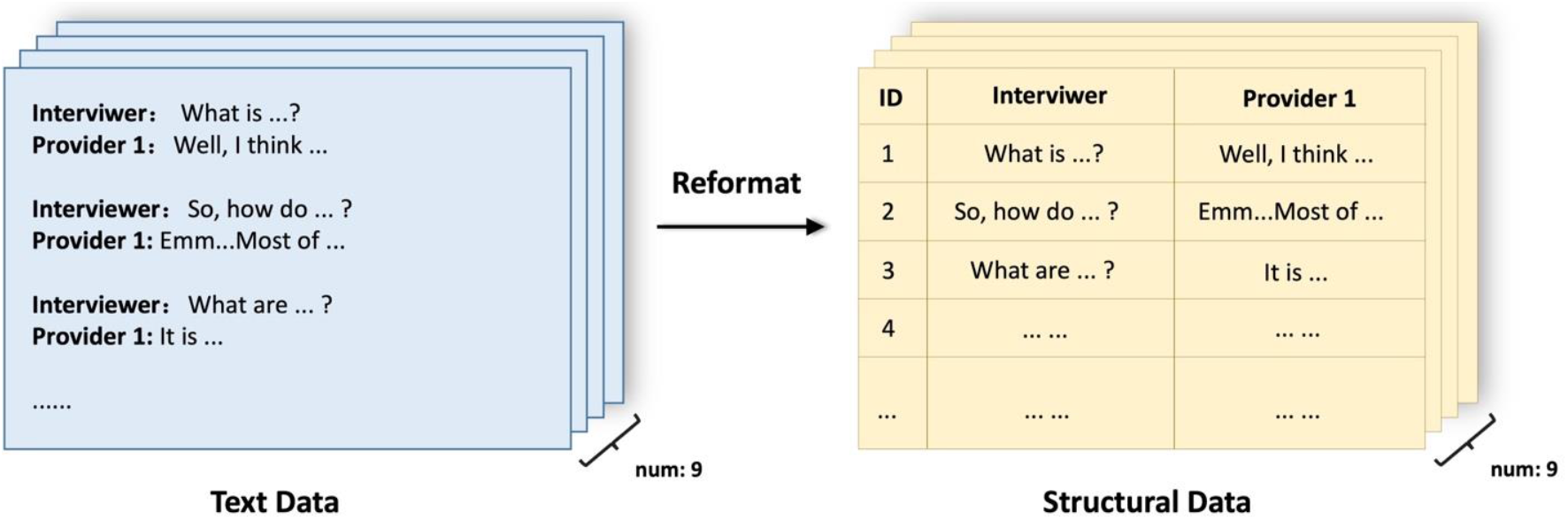
Reformatting raw text data into structured computer-readable data. Each interviewers’ question is matched with its corresponding participants’ answer as a dialogue.

### 2.4. Coding of semi-structured interview with GenAI

In this study, we aimed to investigate the potential of utilizing ChatGPT for thematic analysis in inductive coding for semi-structured interviews. Specifically, through carefully designed instructions and question prompts, ChatGPT generated a set of codes by identifying and summarizing key points mentioned by each participant following thematic analysis approach. Braun and Clarke’s six-phase coding framework widely adopted for thematic analysis of inductive coding was employed in the present study [14]. The framework involves the following steps:

(1) Familiarizing: Read the data thoroughly, transcribe if necessary, and jot down initial ideas.
(2) Generating initial codes: Systematically code features across the dataset, collating relevant data.
(3) Searching for themes: Organize codes into potential themes, gathering relevant data for each theme.
(4) Reviewing themes: Check the application of themes, in relation to the coded extracts and the entire dataset, generating a thematic “map” of the analysis.
(5) Defining and naming themes: Refine themes and the overall narrative, generating clear definitions and names.
(6) Producing the report: Conduct the final analysis, select compelling examples, relate the analysis to the research question and literature, and produce a scholarly report.

In this study, we used ChatGPT 4 to simulate the thematic analysis procedure from the second step to the fourth step. The specific methods included *dialogue filtering, code generation*, and *code aggregation*, which are illustrated in Figure 2. During the development and testing of the proposed workflow, we first interacted with the ChatGPT 4 model through its web-based chatbot interface. This allowed us to iteratively refine prompts utilized for analysis and roughly evaluate model performance in generating appropriate codes. Once the prompt structure was established, we leveraged the ChatGPT API to automate the inductive coding process at scale.

**Figure 2.**
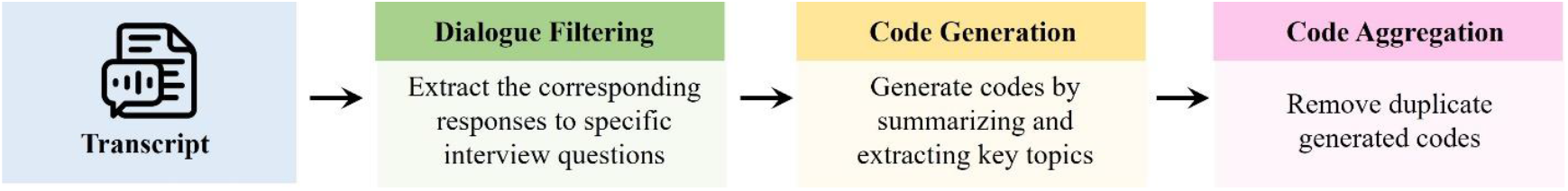
The workflow of using ChatGPT for inductive coding.

#### 2.4.1. Dialogue Filtering

In-depth interviews often feature a deep, conversational nature where participants might address supporting points for one question while responding to another, either unintentionally or to reinforce their viewpoints. As a result, the same perspective might be repeated across different responses and intersect with multiple interview questions. Given this complexity, we performed *dialogue filtering* to locate participants’ responses to specific questions. We first instructed ChatGPT to determine whether a specific question was asked during the interview. When the question was identified, ChatGPT provided the dialogue ID, allowing researchers to observe participants’ responses to the question. This step ensured that only relevant sections of the dialogue are further coded, enhancing the accuracy and efficiency of the analysis. The prompt template used for dialogue filtering was illustrated in Figure 3. The prompt instructed ChatGPT to identify if a specific interview question is present within the dialogue and, if so, to return the ID of the relevant dialogue. If no relevant content was found, it returned a 0. By doing so, for each question, we identified all relevant dialogue from each participant for each question.

**Figure 3.**
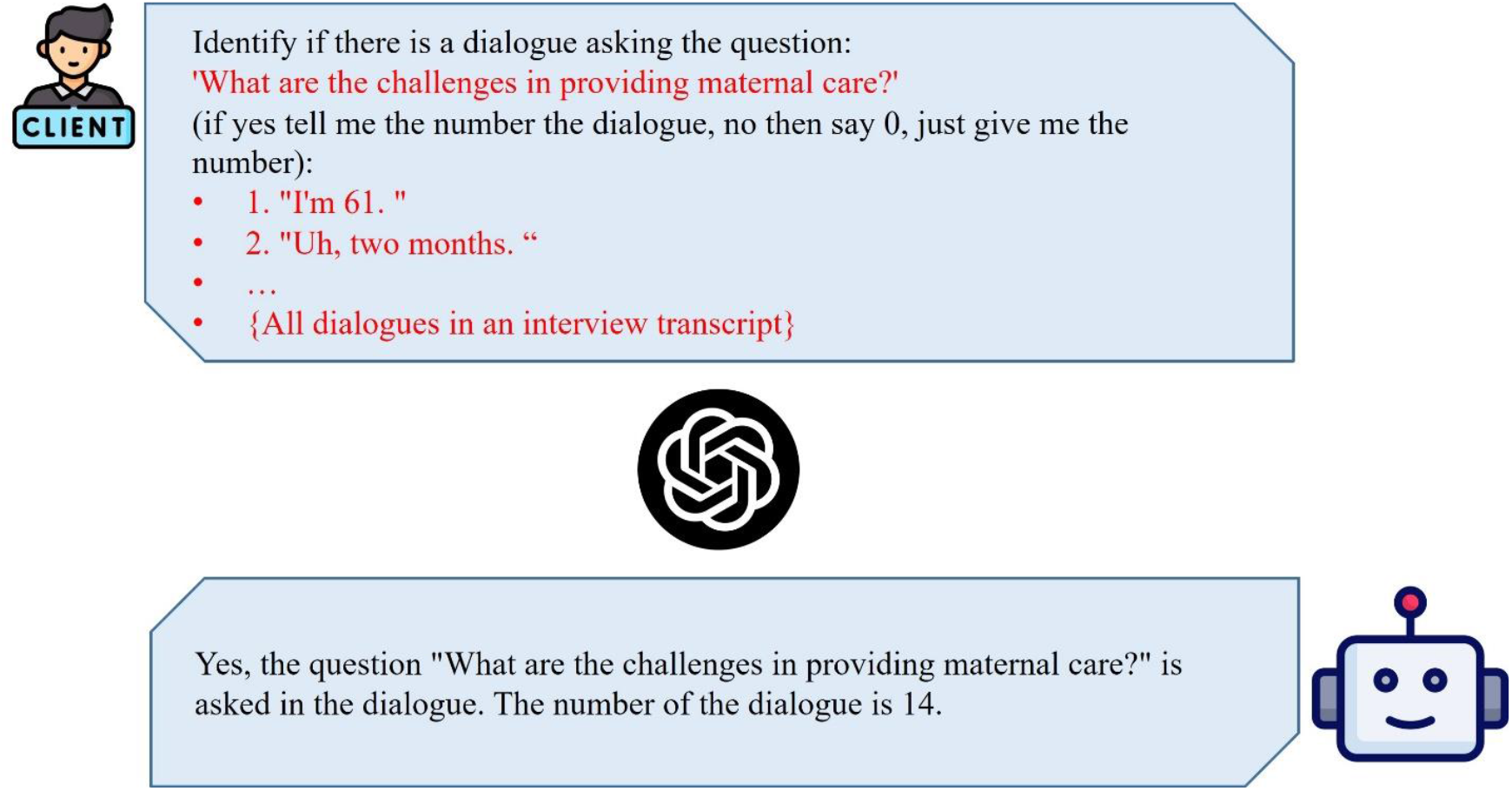
The prompt template used for dialogue filtering. If the given question is asked in the interview transcript, returns the row number of the dialogue, else then returns 0.

#### 2.4.2. Code Generation

After identifying the dialogues for each question, we instructed ChatGPT to perform *code generation* by summarizing and extracting key topics from the dialogues. Figure 4 illustrates the prompt template used for generating codes. In this phase, responses to specific questions from all interview transcripts were collected. ChatGPT was then tasked to extract and generate codes from the collected responses. This process enabled the identification of themes and topics across participants’ responses.

**Figure 4.**
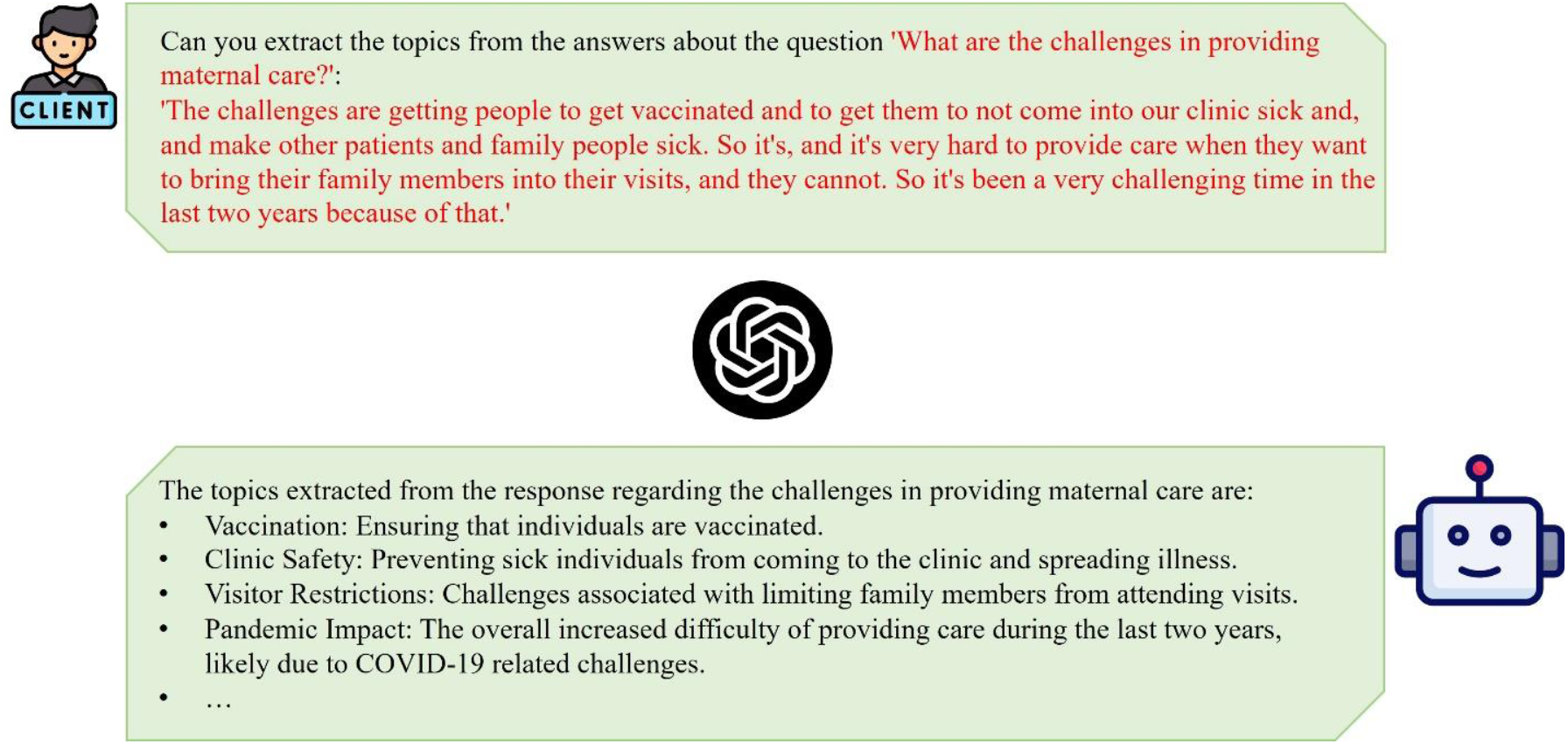
The prompt template used for code generation.

#### 2.3.2. Code Aggregation

After GenAI generated a set of codes reflecting the key topics in response to each question, we further aggregated these codes into categories and developed themes. The first task was to merge codes with similar semantic meanings. While ChatGPT generated the most fitting code for each participant’s response, slight variations in expressions resulted in many repetitive codes that were useless for analysis and required manual aggregation. The second task was to synthesize codes by their conceptual meanings. The codes were combined and grouped into categories based on their definitions and scope in the conceptual system. The process of *code aggregation* ensured that the extracted codes were concise and useful for identifying themes in further qualitative data analysis.

### 2.5. Evaluation

Reviewing the generated and aggregated codes was crucial to ensure the accuracy and applicability of the generated themes. Although ChatGPT is often efficient at identifying general themes present in dialogue, it may lack domain-specific knowledge. Thus, two evaluation approaches were performed to assess the validity of the generated codes: a human-centered approach and a machine-based approach. It is important to emphasize that the purpose of presenting these evaluation methods is not to position one as superior to the other. Rather, our goal is to assess and characterize the performance of ChatGPT through both lenses, recognizing that each approach has its own advantages and limitations.

#### Human-centered approach

We engaged domain experts in maternal health, with relevant background knowledge in manual coding of the interview transcripts. One faculty member and one doctoral student in public health, both with extensive experiences in qualitative studies, independently developed the codes after reviewing all the transcripts. The disagreements were sufficiently discussed and resolved before finalizing codes assigned to each participants’ response. The domain experts reviewed the ChatGPT-generated codes and compared them with the human-generated codes. If the ChatGPT-generated codes were the same and/or aligned with the human-generated codes, they were marked as “accepted”. Two domain experts independently marked the acceptance of each code and discussed and resolved the disagreement. The accuracy rate for each response to a question was the number of “accepted” codes divided by the total number of codes generated by ChatGPT.

#### Machine-based metric approach

We also compared the human-generated codes and ChatGPT-generated codes and quantified the differences by assessing their semantic similarity. The underlying hypothesis is that, if ChatGPT performs effectively, the semantic content of its generated codes should closely resemble that of human-generated codes. In particular, we utilized a cutting-edge text embedding method, Bidirectional Encoder Representations from Transformers (BERT), to convert the codes into high-dimensional vectors [37]. No preprocessing was conducted since the codes (whether generated by humans or ChatGPT) consisted of only a few words or paraphrases. These high-dimensional feature vectors captured the semantic meanings of the input codes. We calculated the cosine similarity between the two high-dimensional vectors using the following formula.

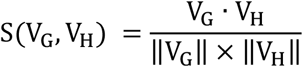

V_G_ represents the high-dimensional vector of a GenAI-generated code, V_H_ represents the high-dimensional vector of a human-defined code, S(V_G_, V_H_) represents the cosine similarity of V_G_ and V_H_. S(V_G_, V_H_) ranges from 0 to 1, and the higher the S(V_G_, V_H_), the more similar the two codes.

## 4. Results

### 4.1 Inductive Coding Performance

We first show several examples of the codes generated by ChatGPT. Table 2 lists the ChatGPT-generated codes for three example questions. Each question was coded based on the participant’s direct responses to the questions.

**Table 2.**
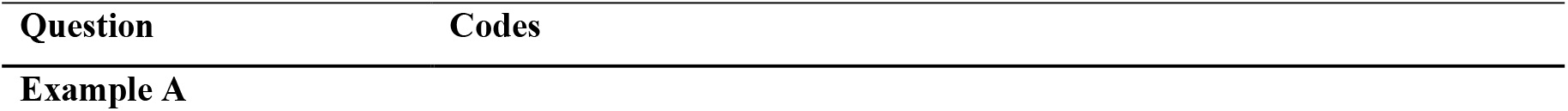
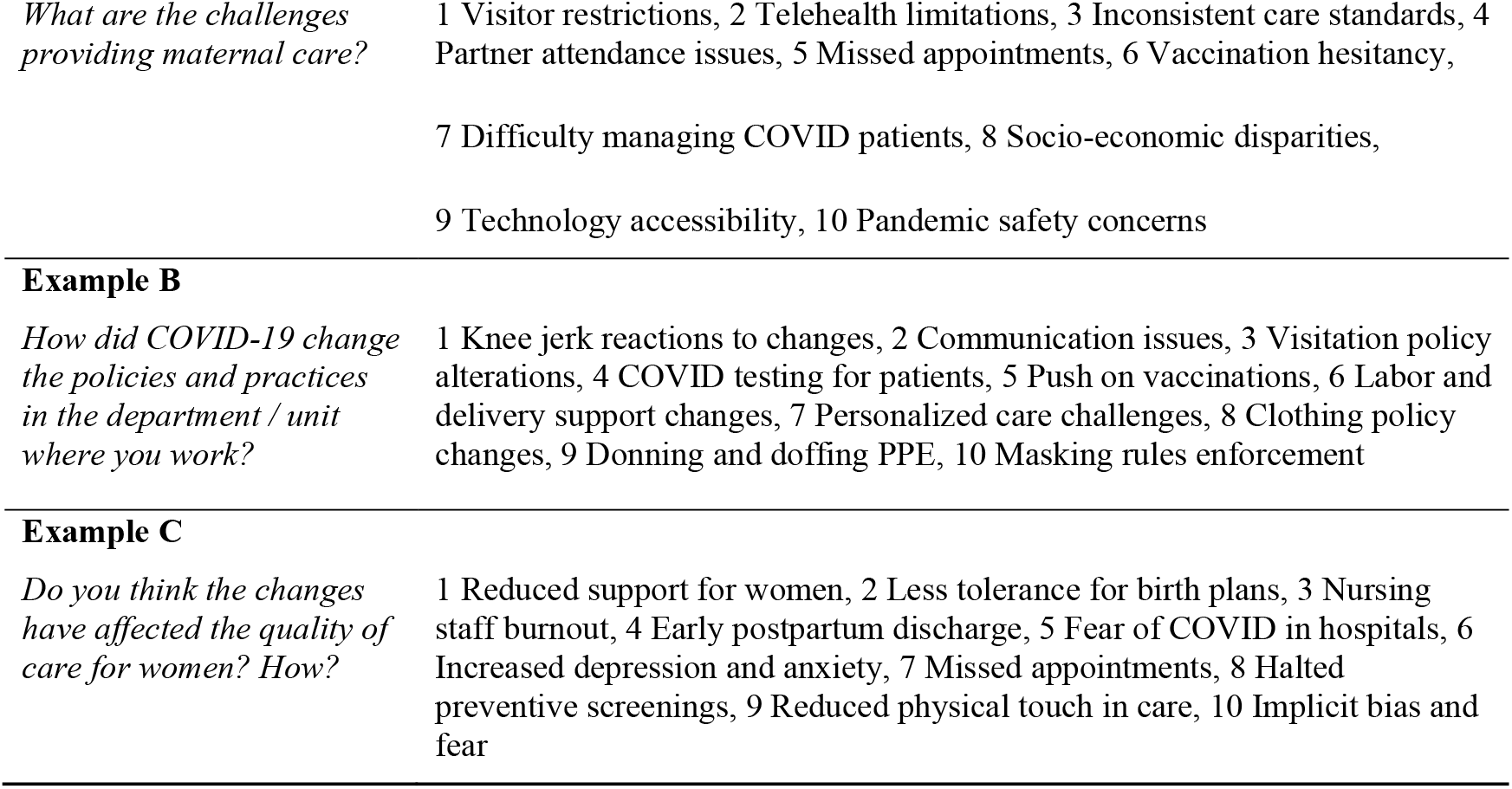
Three example questions and their corresponding codes created by ChatGPT.

| Question | Codes |
| --- | --- |
| Example A |  |
| *What are the challenges providing maternal care?* | 1 Visitor restrictions, 2 Telehealth limitations, 3 Inconsistent care standards, 4 Partner attendance issues, 5 Missed appointments, 6 Vaccination hesitancy, 7 Difficulty managing COVID patients, 8 Socio-economic disparities, 9 Technology accessibility, 10 Pandemic safety concerns |
| **Example B** |  |
| *How did COVID-19 change the policies and practices in the department / unit where you work?* | 1 Knee jerk reactions to changes, 2 Communication issues, 3 Visitation policy alterations, 4 COVID testing for patients, 5 Push on vaccinations, 6 Labor and delivery support changes, 7 Personalized care challenges, 8 Clothing policy changes, 9 Donning and doffing PPE, 10 Masking rules enforcement |
| **Example C** |  |
| *Do you think the changes have affected the quality of care for women? How?* | 1 Reduced support for women, 2 Less tolerance for birth plans, 3 Nursing staff burnout, 4 Early postpartum discharge, 5 Fear of COVID in hospitals, 6 Increased depression and anxiety, 7 Missed appointments, 8 Halted preventive screenings, 9 Reduced physical touch in care, 10 Implicit bias and fear |

Overall, the codes created from the text are relevant and demonstrate alignment—indicating that ChatGPT has provided relatively meaningful results. For instance, the codes generated for the question about challenges in providing maternal care (Example A) cover a broad range of relevant issues including objective (e.g., visitor restrictions), personal (e.g., vaccination hesitancy), and social factors (e.g., socio-economic disparities). These results indicate that ChatGPT has successfully identified and categorized key themes from the participants’ responses. On average, ChatGPT generated approximately 10 codes per interview question, with each code containing an average of five words. Notably, when input the same question into ChatGPT for multiple times, the generated outcomes might be inconsistent. Thus, we perform an assessment to see if the generated results show similar patterns. We leveraged the BERT and computed the similarity among generated codes. The results revealed that the codes generated have a high similarity score of over 0.95, indicating that ChatGPT’s outputs are relatively stable across repeated runs.

ChatGPT’s performance in inductive coding was quantitatively evaluated by accuracy rate of codes for responses to each question. Figure 5 presents the number of codes generated by GenAI for each question, the number of “accepted” codes after manual review, and the corresponding accuracy rates. The overall accuracy rate was observed to be 85.15% across all questions. For individual questions, the accuracy rates ranged from a maximum of 100% to a minimum of 66.67%. Notably, 11 questions had coding accuracy rates equal to or greater than 90%, representing more than half of the total number of questions, with 8 questions achieving a coding accuracy rate of 100%. Only three questions had coding accuracy rates less than 80%. After reviewing these three questions, we found that the responses are highly descriptive and ChatGPT tended to extract more generalized information from these detailed responses. For example, in coding Question 6, “Do you think the changes have affected maternal health outcomes? How?”, ChatGPT provided the broad summary code of “Changes affecting maternal health.” For Question 10, which asks, “How would you like to describe your clients’ psychological conditions during the pregnancy in general?”, ChatGPT provided summarized codes like “Patients’ health and baby’s health concerns”, while human-generated codes were more specific, such as “Psych,” “Anxiety”, and “Anger”. This finding indicated that a small portion of questions did not achieve high accuracy in coding. Overall, the inductive coding results are reliable and ChatGPT demonstrated promise in providing support for qualitative coding.

**Figure 5.**
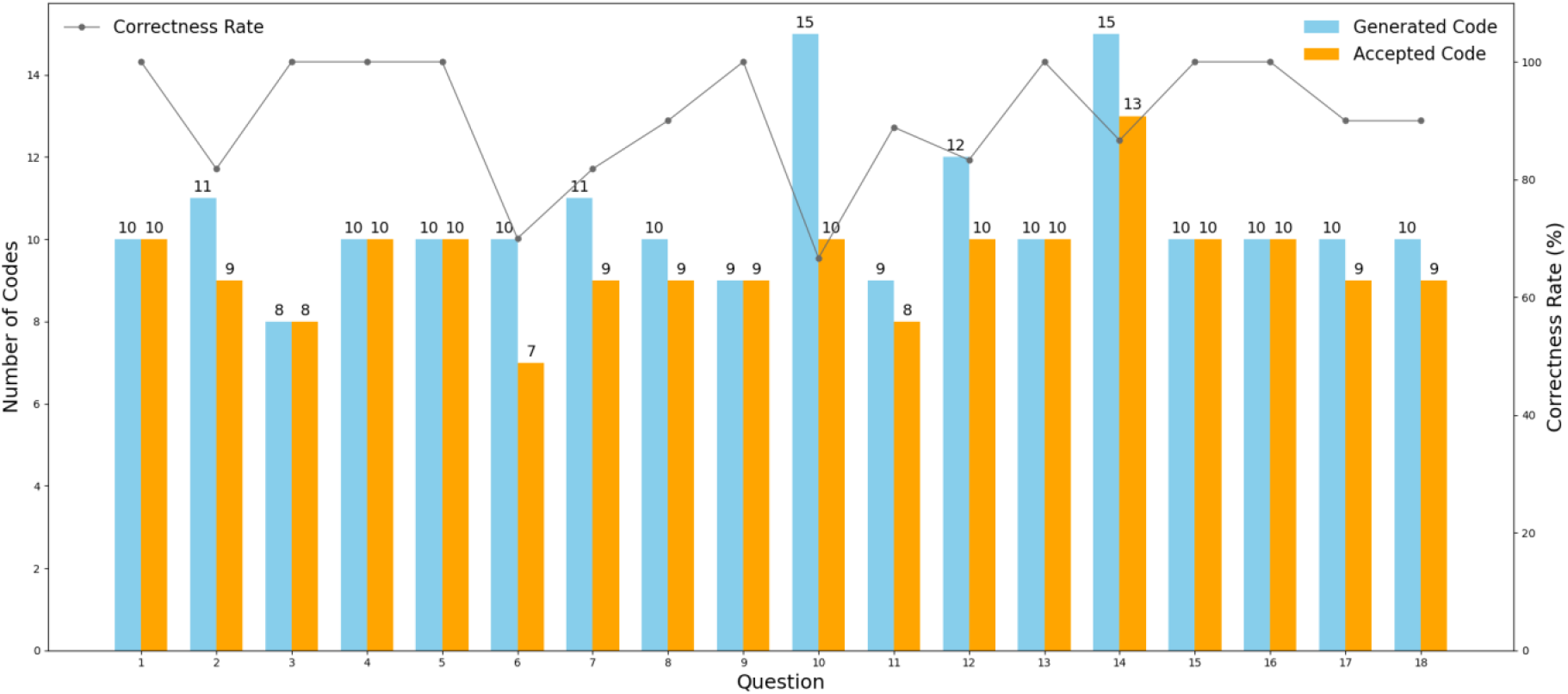
The number of generated codes, the number of “accepted” codes after manual review, and the accuracy rate for each question.

Moreover, we compared the ChatGPT-generated codes and human-generated codes for each question using the text semantic similarity score. Figure 6 shows the cosine similarity between the high-dimensional vectors of the two sets of texts. The highest similarity score was 0.879, with an overall similarity of 0.795. It should be noted that there is no established threshold for determining semantic similarity between two groups of codes. Thus, to provide greater interpretability of these results, we present three sample codes, as illustrated in Table 3 with high, medium, and low semantic similarity scores. This indicates that the semantic meaning of the codes generated by ChatGPT are very similar to those by human coders, illustrating the proficiency of using ChatGPT for the inductive coding of interview texts.

**Table 3.**
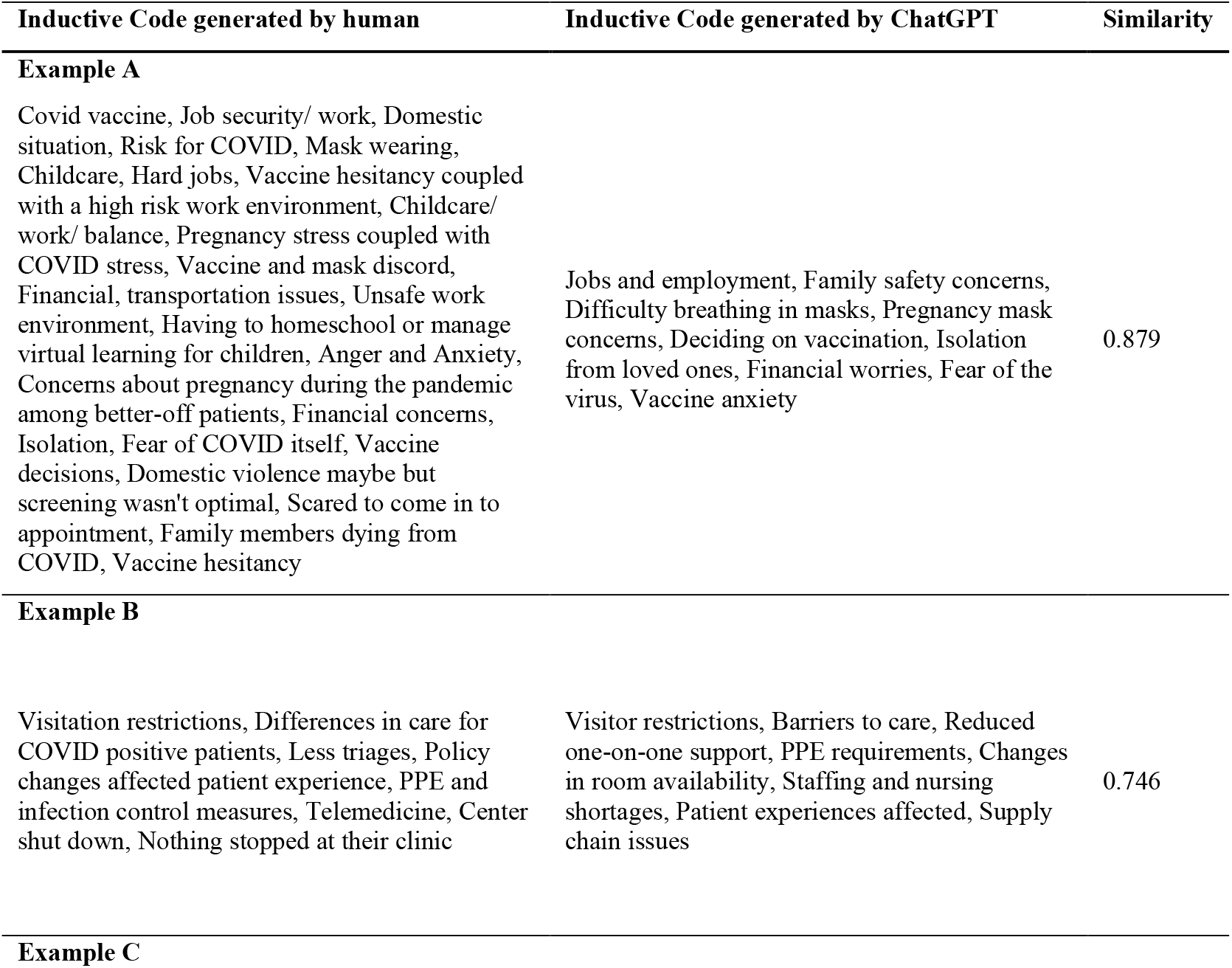
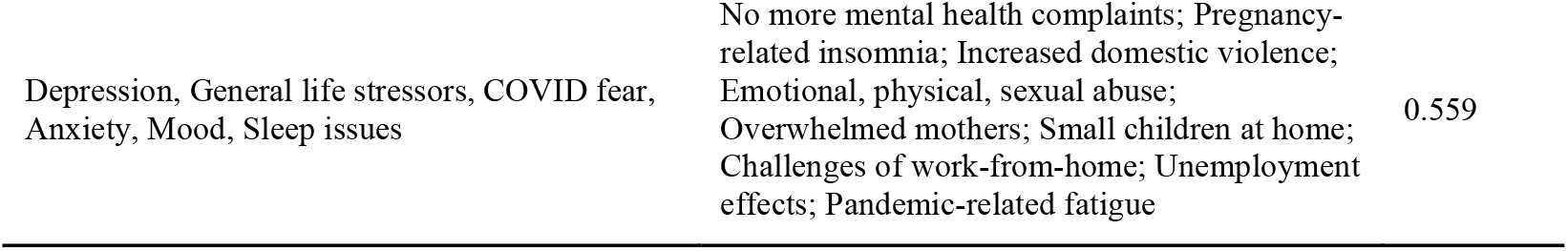
Similarity of inductive codes generated by human-coders and ChatGPT.

**Figure 6.**
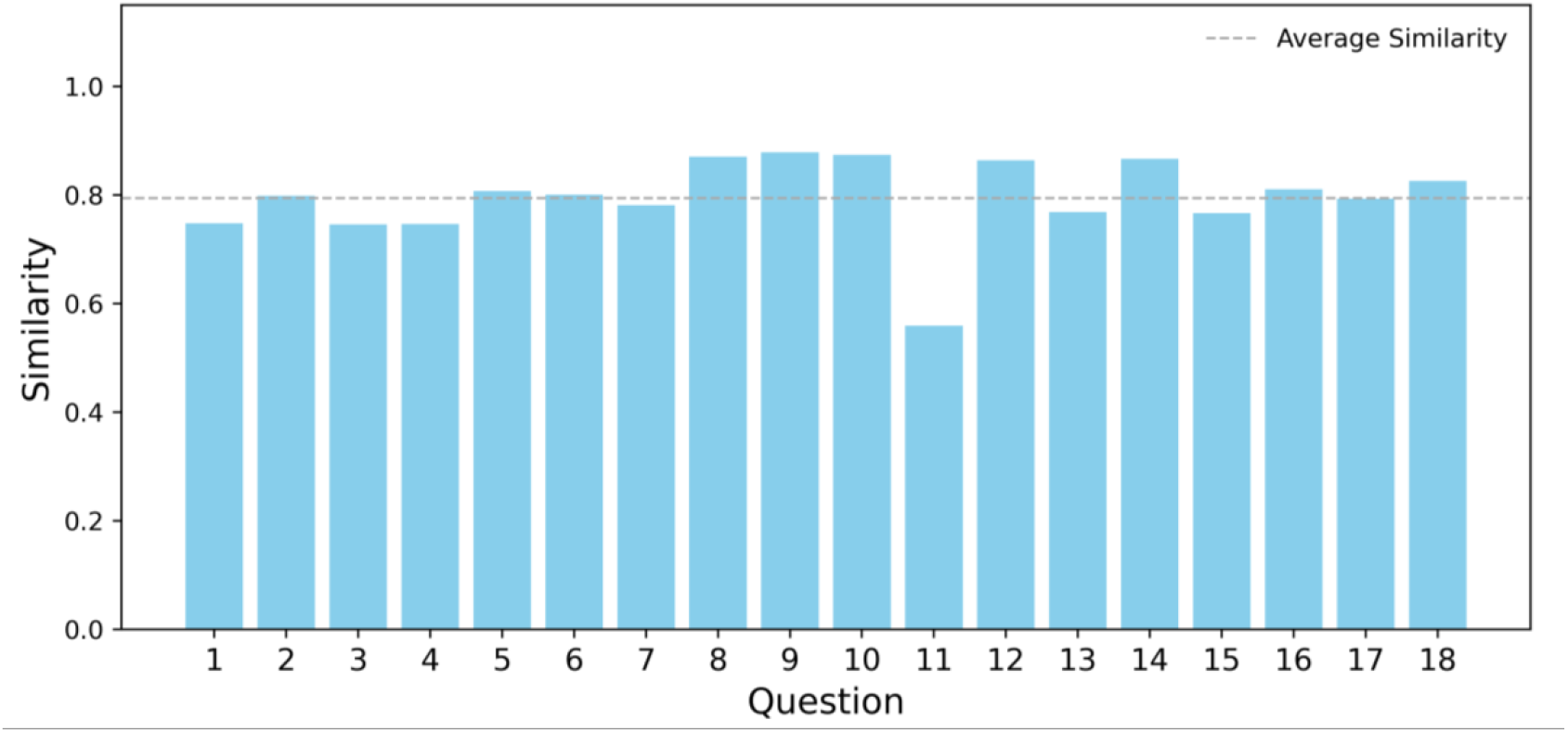
Text similarity between the ChatGPT-generated codes and human-generated codes for each question. The x-axis represents text similarity, and the y-axis represents the question number.

### 4.2 Time efficiency

In addition to demonstrating accuracy, the GenAI-based coding approach has also shown significant time efficiency compared to traditional methods. The total estimated time spent by human coders for inductive coding was approximately 30.5 hours, of which 4.5 hours were spent getting familiar with the 9 transcripts and 11.25 hours were spent in generating and assigning codes to the dialogues.

The usage of ChatGPT was found to significantly reduce the time required to complete inductive coding. The time expense of GenAI-based coding primarily reflects two activities: *setup* and *coding*. The first is the time spent setting up the environment, including data preparation for ChatGPT coding, designing appropriate prompts and reformatting the transcript files. The second is the time required for the actual coding, by ChatGPT. The first part, setting up the environment, requires most of the time in the work pipeline, around 5 hours. In particular, a human coder takes 3 hours to reformat the original transcript into Excel to prepare the materials for use in the GenAI coding. Then, it takes around 2 hours to test different prompts and reformat the transcript files to be fed into GenAI. Once the setup is complete, the actual coding process by GenAI is much faster, taking only around 20 minutes to generate and assign codes following the inductive approach.

It should be noted that additional human-centered tasks are still necessary with current GenAI techniques. For example, these methods require domain experts to review the coding results and assess the quality of how each transcript was coded, which results in additional time costs. However, with the development of more robust and advanced GenAI models in the future, these review steps may be simplified, and the process may be shortened. Consequently, the total time required for GenAI-based coding might be reduced to just 19% of the time required for traditional coding methods.

## 5 Discussion

These results of using ChatGPT for inductive coding highlight the feasibility of GenAI in qualitative studies that use a post-positivist approach to Braun and Clarke’s thematic analysis [14]. ChatGPT achieved relatively high-performance inductive coding. Traditionally, coding interview transcripts manually is a time-consuming process. Using GenAI in coding can greatly shorten this process and maintain relatively high accuracy. Thus, it could enhance efficiency in qualitative research. The findings from our study provide preliminary evidence for both opportunities and challenges of using GenAI in qualitative data coding.

### 5.1 Opportunities brought by GenAI for qualitative study

The emergence of GenAI technology provides unprecedented opportunities to enhance coding of qualitative data to support thematic analysis.

#### User friendly and accessible

GenAI is easily accessible in terms of cost and ease of use, accommodating users with varying levels of technical expertise. This accessibility allows for broad applicability across various research domains without requiring extensive prior knowledge or domain-specific input. For example, ChatGPT offers both web-based chatbot user interface and APIs. Thus, users could interact with it like human conversations. Also, ChatGPT APIs facilitate integration of GenAI into custom workflows and enable large-scale, automated processing of textual data. The advanced natural language understanding ability makes GenAI automatically screen interview transcripts and identify specific questions or topics related to a given question. When designing prompts in ChatGPT, there is no need to input a large training dataset. Even when using a small number of sample dialogues, ChatGPT promisingly achieved high levels of accuracy in identifying and categorizing topics in the data. In our study, although the main research topic is related to maternal health, ChatGPT successfully identified discussions related to cannabis, opioid, and alcohol use from the participants’ responses, showing its flexibility across different public health areas. This suggests GenAI can be effectively applied across public health research.

#### Multi-level semantic structure identification

GenAI demonstrates an ability to identify multi-level semantic structures when coding, which is well represented in the code aggregation process. ChatGPT can generate fine-resolution codes, and merge those with similar semantic meanings into broader categories. This granularity of analysis provides a more detailed understanding of qualitative data, enabling researchers to capture the complexity and diversity of participants’ perspectives. Traditionally, qualitative researchers must manually merge and aggregate codes at different levels. With GenAI, subtopics can be automatically grouped under their main topics, enhancing the overall coherence of the analysis.

#### Scalability and efficiency in large dataset

GenAI excels at handling large datasets, providing a scalable solution for the analysis based on large datasets. Analyzing large datasets is often a challenge due to time and human resource limitations. Once the environment setup is completed, GenAI can process transcripts in parallel by leveraging its Application Programming Interfaces (APIs), which can dramatically improve coding efficiency. Therefore, GenAI can serve as a tool, completing most of the initial coding tasks, thereby allowing human coders and experts to expend their efforts on the careful review and discussion of outputs as well as the applicability, interpretation, and dissemination of the findings. Thus, collaboration between GenAI and human coders can potentially shift the time required for qualitative data analysis while preserving the quality of the data analysis.

### 5.2 Challenges and Concerns When Using GenAI

Despite the promising potential of GenAI for coding in qualitative studies, several challenges and concerns need careful consideration, including ***inaccuracy, systematic biases, and privacy issues***. The inaccuracy issues in GenAI-based coding stems from two aspects. First, the lack of maternity knowledge, due to the sensitivity nature of maternal health research, may result in inaccurate results. We observed that ChatGPT tends to produce more general codes than human coders and lacks the domain-specific terminology and professional knowledge unique to maternal experts. For example, for Question 10, “How would you like to describe your clients’ psychological conditions during the pregnancy in general?”, one of the codes generated by ChatGPT was “Post-traumatic stress from having COVID,” while the human-generated code was “Some form of PTSD symptoms in patients who were really sick with COVID,” which used more professional medical terminology and provided a more detailed description of the psychological issues described. This discrepancy may be attributed to ChatGPT being trained on a general corpus with limited exposure to public health knowledge, especially maternal health knowledge. Also, maternal health data is often highly sensitive, hinders the ability of ChatGPT in learning maternity knowledge. The implications of this are that qualitative studies seeking to use GenAI-based coding to assist within their data analysis will need to consider selecting a model that can be trained on their public health topic of interest. Second, the “hallucination” phenomena in commonly observed in current GenAI models. Given that current GenAI models, such as ChatGPT, are trained on a general corpus, rather than specific domain knowledge, GenAI can produce inaccurate content and false information. Therefore, these hallucinations further demonstrate the necessity of human coders and experts to carefully review all outputs generated by GenAI during the coding process.

Additionally, systematic biases are rooted in representation issues when training datasets. As the behind-the-scenes generation process of GenAI is inaccessible to prompt engineers, it is difficult to quantify errors and debug them. The datasets used to train GenAI are not readily available to users and may underrepresent certain minority population groups, thereby overlooking their lived experiences, socio-linguistic differences, and potentially leading to the amplification or replication of existing social biases [38]. Being able to capture and understand these nuances are essential within qualitative research, as it is not possible to understand a phenomenon of interest without doing so. Since, thematic analysis looks at patterns across a dataset, researchers conducting latent level analysis using GenAI-developed codes are particularly vulnerable to unknowingly drawing conclusions that are decontextualized and incorrect [14]. Anticipating and preventing these instances of error are vital as studies using GenAI-developed codes could inform and influence clinical practices and public health policies.

Moreover, using GenAI systems, such as ChatGPT, poses privacy concerns, particularly regarding the handling of sensitive or personal interview data. Current GenAI systems may retain and use input data for further model training, which could lead to misuse or sharing of private information. Researchers must sufficiently assess potential ethical issues before entering sensitive information into GenAI models, adhere strictly to IRB guidelines to ensure compliance with data protection protocols. Implementing robust data security measures, including data de-identification, and stringent access controls, can mitigate privacy risks. In addition, potential technical solutions may help address this issue in the future. The development of privacy-preserving machine learning techniques, such as differential privacy and secure multi-party computation, can enhance data protection without compromising the utility of GenAI models in qualitative research. Furthermore, techniques such as machine unlearning may protect sensitive data by preventing models from retaining or memorizing sensitive input data, thereby contributing to more secure and ethically use of large language models. Ultimately, ensuring the responsible use of GenAI in public health research necessitates a multidisciplinary approach that integrates technological advancement, regulatory compliance, and further discussions on ethics. This paper highlights such importance and advocates for more interdisciplinary collaborations to harness the benefits of GenAI and maintain the highest standards of data integrity, participant confidentiality, and rigor in qualitative methodology.

### 5.4 Limitations and Future Directions

This study has several limitations that need to consider. First, we analyzed transcripts from individual semi-structured interviews with maternity care providers conducted during the COVID-19 pandemic. The specificity of this dataset may limit the transferability of our findings to other interview formats (e.g., structured interviews, unstructured interviews, or focus groups) and different research contexts (e.g., different geographic regions, various healthcare settings, or alternative qualitative analytical approaches). Future studies should apply the computational workflow across a wider range of datasets and research contexts to showcase its adaptability and potential in qualitative research.

Second, in the current study, we tested only one GenAI model (i.e., ChatGPT). Variability in performance, accuracy, and interpretability across different GenAI models (e.g., Gemini, DeepSeek) remains an open question. Future studies may explore the potential to incorporate more advanced GenAI approaches such as agent-based GenAI methods, or different GenAI models (e.g., commercial vs. open-sourced models). Comparing different models will provide insights into their relative strengths, weaknesses, and applicability to qualitative research.

Third, in our study, we used accuracy rate based on two human coders’ judgment to assess the credibility of AI-generated codes against the human generated codes within our dataset. Within qualitative research using thematic analysis, many tools exist to assess the accuracy of manually coded transcripts (e.g., reflexivity journals, triangulation, internal audits, prolonged engagement, member checks, thick descriptions), with the selection each tool being dependent on which qualitative research paradigm a study is situated within. In addition, the potential for anchoring bias among human coders must be considered during the evaluation of AI-generated codes. When reviewing outputs produced by large language models (LLMs), human evaluators may be more inclined to accept the suggested codes rather than critically proposing or independently generating alternative codes. Therefore, future research needs to explore the best practices for assessing the quality and validity of qualitative coding when incorporating LLMs.

Fourth, we did not train our LLMs on codes relevant to maternal health. This limitation may have resulted in the model overlooking critical domain-specific nuances in the coding process. Future research is needed to assess the degree to which coding quality changes when the LLMs are trained versus untrained.

## 6 Conclusion

In summary, we proposed a computational framework for coding qualitative data for thematic analysis by leveraging advanced GenAI methods. Our findings demonstrated the potential of GenAI (i.e., ChatGPT 4) in automating the inductive coding processes, thereby enhancing efficiency within qualitative studies using Braun and Clarke (2006)’s approach to thematic analysis. In our case study on maternal health, ChatGPT could generate meaningful codes from the data to uncover key themes and topics from the semi-interview transcripts. Our evaluation showed a relatively high overall accuracy of GenAI-generated codes and a significant reduction in the time spent on coding processes. These findings suggest that GenAI can effectively support qualitative coding, provide reliable results, and offer substantial benefits for various public health studies. Despite its promise, we have also noted the challenges in applying GenAI in qualitative research, including inaccuracies, systematic biases, and privacy issues. Therefore, it is crucial to incorporate more domain knowledge into GenAI and have domain experts carefully review the results of GenAI-based coding given the current technologies. With advancement in AI technologies, we believe qualitative research will continue to benefit from these cutting-edge methods.

## Data Availability

All data produced in the present study are available upon reasonable request to the authors

## Acknowledgement

The authors would like to thank Camryn Garrett and Ariele N’Diaye at the University of South Carolina who helped us preprocess the maternal dataset and proofread the manuscript. Y.K. acknowledges the funding support provided by the Population Research Center Seed Grant, The University of Texas at Austin. M.L. acknowledges the funding support provided by NIH grant #R01AI127203-05S2. S.Q. acknowledges the funding support provided by NIH grant#R01AI174892. Any opinions, findings, conclusions, or recommendations expressed in this material are those of the author(s) and do not necessarily reflect the views of the National Institutes of Health (NIH)

